# Machine learning based prediction of prolonged duration of mechanical ventilation incorporating medication data

**DOI:** 10.1101/2023.09.18.23295724

**Authors:** Andrea Sikora, Bokai Zhao, Yanlei Kong, Brian Murray, Ye Shen

**Affiliations:** University of Georgia College of Pharmacy, Department of Clinical and Administrative Pharmacy, Augusta, GA, USA 1120 15th Street, HM-118 | Augusta, GA 30912 |; University of Georgia College of Public Health, Epidemiology & Biostatistics, Athens, GA, USA; Renmin University of China, School of Statistics, Beijing, China; University of North Carolina Medical Center, Department of Pharmacy, Chapel Hill, NC, USA; University of Georgia University of Georgia College of Public Health, Epidemiology & Biostatistics, Athens, GA, USA

## Abstract

**Rationale:** Duration of mechanical ventilation is associated with adverse outcomes in critically ill patients and increased use of resources. The increasing complexity of medication regimens has been associated with increased mortality, length of stay, and fluid overload but has never been studied specifically in the setting of mechanical ventilation.

**Objective:** The purpose of this analysis was to develop prediction models for mechanical ventilation duration to test the hypothesis that incorporating medication data may improve model performance.

**Methods:** This was a retrospective cohort study of adults admitted to the ICU and undergoing mechanical ventilation for longer than 24 hours from October 2015 to October2020. Patients were excluded if it was not their index ICU admission or if the patient was placed on comfort care in the first 24 hours of admission. Relevant patient characteristics including age, sex, body mass index, admission diagnosis, morbidities, vital signs measurements, severity of illness, medication regimen complexity as measured by the MRC-ICU, and medical treatments before intubation were collected. The primary outcome was area under the receiver operating characteristic (AUROC) of prediction models for prolonged mechanical ventilation (defined as greater than 5 days). Both logistic regression and supervised learning techniques including XGBoost, Random Forest, and Support Vector Machine were used to develop prediction models.

**Results:** The 318 patients [age 59.9 (SD 16.9), female 39.3%, medical 28.6%] had mean 24-hour MRC-ICU score of 21.3 (10.5), mean APACHE II score of 21.0 (5.4), mean SOFA score of 9.9 (3.3), and ICU mortality rate of 22.6% (n=72). The strongest performing logistic model was the base model with MRC-ICU added, with AUROC of 0.72, positive predictive value (PPV) of 0.83, and negative prediction value (NPV) of 0.92. The strongest overall model was Random Forest with an AUROC of 0.78, a PPV of 0.53, and NPV of 0.90. Feature importance analysis using support vector machine and Random Forest revealed severity of illness scores and medication related data were the most important predictors.

**Conclusions:** Medication regimen complexity is significantly associated with prolonged duration of mechanical ventilation in critically ill patients, and prediction models incorporating medication information showed modest improvement in this prediction.

## Introduction

Mechanical ventilation (MV) is a frequently encountered supportive care modality in critically ill patients. (1) While life-saving, mechanical ventilation predisposes patients to notable sequalae that can adversely affect patient-centered outcomes including ventilator induced lung injury (VILI), infection, intensive care unit (ICU) acquired muscle weakness, etc. that make the decision to intubate, daily evaluation of extubation potential, and need for tracheostomy essential bedside assessments. (2) Moreover, the risk of these MV-related complications increases as duration of mechanical ventilation increases, and given that over 750,000 patients are supported by MV and approximately 5-25% of those patients in a mixed ICU require MV for greater than 5 days, prediction of patients at most risk becomes increasingly relevant. (3–5) The ability to identify those patients more likely to require prolonged mechanical ventilation (PMV) may alter clinical decision-making, including ventilation or medication management strategies, that may ultimately reduce the duration of therapy.

Previous studies that developed prediction models for PMV have included primarily laboratory and vital sign information (e.g., serum creatinine, acidosis, bicarbonate, etc.); however, mechanical ventilation cannot be viewed as an isolated intervention but is deeply intertwined with a patient’s overall clinical status and associated pharmacotherapy regimen. (1,5–11) Indeed, associated pharmacotherapy regimens are complex with over 30% of patients in the ICU setting having greater than 20 medications prescribed. (12,13) This same study showed that 70% of patients had more than 13 medications at any given point. (12,13) Further, comprehensive medication management strategies aimed at optimizing drug selection (e.g., preferring medications with lower risk of delirium) have shown reductions in duration of mechanical ventilation. (14,15)

Given that medications play a significant role in mechanical ventilation management, quantifying patient-specific, medication-related data is a potentially important prediction variable that has not been well explored. Previous studies have observed associations between medication regimen complexity-intensive care unit (MRC-ICU) score and other relevant outcomes including mortality, length of stay, and fluid overload. (16–24) Moreover, use of machine learning techniques in combination with MRC-ICU appears to improve prediction and has shown utility in a variety of ICU prediction questions. (20,25)

In this study, we aimed to employ both traditional and machine learning techniques to develop and validate PMV prediction models with the goal of identifying the most useful predictors at the bedside. We hypothesized that advanced machine learning techniques may be useful to identify the most important clinical factors that can differentiate between patients with high versus low risk of PMV. Additionally, we aimed to test the hypothesis that the addition of medication related variables would enhance prediction of PMV.

## Methods

### Source of data

Patient data were gathered from the Carolina Data Warehouse, which houses Epic^®^ electronic health record (EHR) data from the University of North Carolina Health System. The protocol for this study was reviewed and approved by the University of Georgia Institutional Review Board (approval number: (Project00001541). Due to the retrospective, observational design, waivers of informed consent and HIPAA authorization were granted.

This was a retrospective cohort study of adults aged 18 years and older with an ICU admission and duration of mechanical ventilation greater than 24 hours between October 2015 and October 2020. Patients were excluded if it was not their index ICU admission or if the patient was placed on comfort care within the first 24 hours of ICU stay. Types of ICUs included medical, surgical, trauma, neurosciences, cardiac, and burn. De-identified data from the first ICU admission per each patient was included.

The primary outcome was the presence of prolonged duration of mechanical ventilation, defined as greater than 5 days. De-identified patient data were collected through a trained data analyst including: 1) baseline demographic data: age, sex, admission to the medical ICU, primary admission diagnosis, and data to calculate the sequential organ failure assessment (SOFA), Acute Physiology, Chronic Health Evaluation (APACHE II) score, and medication regimen complexity-intensive care unit (MRC-ICU) score at 24 hours; 2) data at time of mechanical ventilation: highest FiO_2_ positive end expiratory pressure (PEEP), and minute ventilation in 12-24 hours after intubation, mode of ventilation at initiation, duration of non-invasive respiratory support prior to intubation, arterial blood gas values, and heart rate; 3) patient outcomes: mortality, ICU length of stay, presence of fluid overload, acute kidney injury (AKI), and need for vasoactive medications and/or renal replacement therapy; and 4) medication variables: sedatives, analgesics, vasoactive agents, and diuretics taken before intubation. Additionally, the MRC-ICU was calculated at 3 separate time points: 24 hours after ICU admission, at the time of intubation, and 24 hours after intubation. The MRC-ICU measures medication regimen complexity with each line of 35 discrete line items assigned a weighted value and then summed to create a score for a patient’s regimen at the given time point. (19) For example, a patient would be given 3 points for vancomycin, 1 point for norepinephrine, and 1 point for insulin for a total score of 5.

### Statistical Analysis

Following a literature review of prolonged mechanical ventilation in the ICU, a total of 30 potential predictor variables were identified by investigator consensus to include in each PMV regression model. (5,10,26–33) These variables included the following: *1) ICU baseline:* age, sex, admission to medical ICU, and primary ICU admission diagnosis (cardiac, chronic kidney disease, heart failure, hepatic, pulmonary, sepsis, trauma); *2) 24 hours after ICU admission:* APACHE II and SOFA score (using worst values in the 24 hour period); presence of acute kidney injury; *3) Flowsheet and laboratory values at time of intubation:* duration of oxygen support devices prior to intubation, pulse, arterial blood gas values, and ventilator settings (positive end expiratory pressure (PEEP) and fraction of inspired oxygen (FiO_2_); fluid overload. The fourth category was added to assess the role of medications at 24 hours and included: MRC-ICU score at 24 hours after ICU admission, time of intubation, and 24 hours after intubation, dichotomized MRC-ICU less than or greater than or equal to 10 at the same three time points, and certain medications (bumetanide, clonazepam, dexamethasone, dexmedetomidine, dobutamine, ertapenem, lorazepam, norepinephrine) selected by the investigators following univariate analysis.

Due to the hypothesis-generating nature of this evaluation, no attempt was made to estimate the sample size, and all eligible patients from the available database were included to maximize statistical power of the predictive models developed. Descriptive statistics on the data were calculated before model development. Numerical predictors were summarized by the mean and standard deviation and categorical variables reported count and corresponding proportion of the total population. Clinical characteristics between those patients with and without PMV were compared using either Student *t* test or Chi-square test, as appropriate.

Univariate analysis was performed on baseline variables to detect potential important variables for the basic model and select important baseline medications for predicting prolong MV.

Multiple imputation was applied for missing data. Under the assumption of missing at random, a chained equations approach carried out five imputations. (34) After multiple imputation, each of the five completed datasets was split into training data and testing data using an 85:15 ratio in a cross-validation splitting manner. Subsequently, a logistic model was built to predict PMV on each of the five complete training sets. Parameter estimation were pooled using Rubin’s rule. (35) Corresponding odds ratio (OR) and 95% confidence intervals (CI) of coefficients were reported respectively, which can reflect the influence of each predictor.

### Model development and performance

Multivariable logistic models were developed to predict PMV. First, a benchmark model was derived using logistic regression and investigator selection that included the following variables: duration of oxygen support devices prior to intubation, highest PaCO_2_ and bicarbonate on arterial blood gas in 12-24 hours after intubation, presence of elevated pulse (>110 beats per minute) in 24 hours prior to intubation, presence of fluid overload before intubation (defined as greater than 10% of admission body weight in volume), highest FiO_2_ and PEEP in 12-24 hours after intubation, minute ventilation, and a product of PaCO_2_ and minute ventilation in the first 12-24 hours after intubation.

Then, the effects of inclusion of standard severity of illness scores (SOFA score at 24 hours, APACHE II score at 24 hours) and medication related data including both the MRC-ICU at 24 hours and presence of individual drugs were assessed. To evaluate the influence of the MRC-ICU score on PMV, MRC-ICU as a continuous score and MRC-ICU as a dichotomous variable as < 10 or ≥10 were included separately, with selection of the rest of the variables following this addition. Additional models including all medications taken before intubation and selected medications known to play a significant role in mechanical ventilation were evaluated for their presence at initiation of mechanical ventilation and 24 hours after intubation. For this process, univariate analysis and then least absolute shrinkage and selection operator (LASSO) based logistic model were conducted on the medication dataset for variable screening resulting in two models (base + selected drugs and base + LASSO drugs). All variables were examined, and certain variables were excluded from the models if potential collinearity or mediation effects were present. A full model was fitted with all predictors, and a stepwise regression was conducted. Additionally, variable selection was performed in the MRC-ICU model by mediation analysis. Variables as mediators with significant relationship between MRC-ICU and PMV were removed from the model. The final model was selected based on the best performance.

Figure 1 in the **Supplemental Content** summarizes the full model development procedure. Multiple logistic models with different predictor variable were built including basic variables, basic variables and MRC-ICU, basic variables and important medications selected via LASSO technique, basic variables with SOFA and APACHE II scores, and basic variables with MRC-ICU, SOFA and APACHE II scores at 24 hours. To compare performance of the models, a series of values were calculated including for the models with classification task: area under the receiver operating characteristic (AUROC), positive predictive value (PPV), negative predictive value (NPV), specificity, and sensitivity. To evaluate performance of the developed model, metrics were calculated on test data from each imputed dataset and averaged. A p-value less than 0.05 was used to determine statistical significance for all outcomes. All analyses were performed using *R* (version 4.1.2).

**Figure 1.**
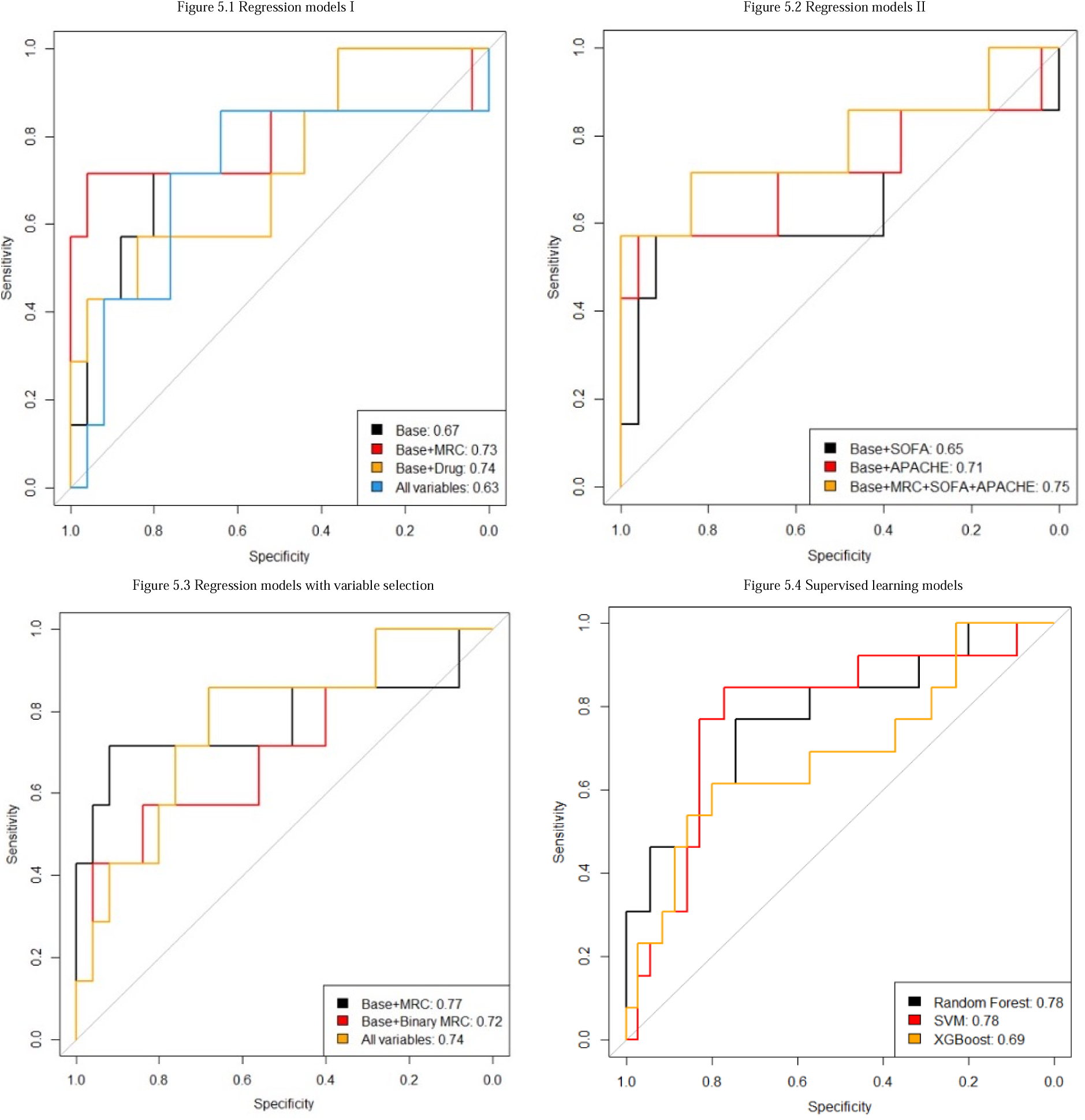
Mean AUROC curve comparing prediction models for prolonged duration of mechanical ventilation.

Random Forest, support vector machine (SVM), and XGBoost were employed for the task of predicting PMV. For these methods, the dataset was imputed and split one time. The mean was used to impute numerical predictors and mode for categorical variable imputation. The dataset was then split into training and test subsets by the ratio of 85:15. Predictions for probability of PMV were made on each of the five imputed testing sets with the corresponding optimal model. Cross-validation was employed for hyperparameters. For Random Forest, two hyperparameters were tuned (number of trees and number of variables randomly sampled as candidates at each split). With the optimal models, the prediction performance on the test dataset was evaluated. For XGBoost, feature importance was measured as the frequency a feature was used in the trees. For Random Forest, feature importance was measured by mean decrease in node impurity. For SVM, feature importance was measured via permutation test from R package ‘vip’. Then, impacts of predictors on the outcome via the evaluation of feature importance and logistic parameter estimation results were compared.

## Results

Of the 318 patients included, 145 patients (45.6%) were over the age of 65, and 125 (39.3%) were female. Of these, 90 (28.3%) had PMV. Overall, the mean APACHE II score was 21.0 ± 5.4, mean SOFA score was 9.9 ± 3.3, and MRC-ICU was 21.3 ± 10.5. The mean length of MV was 5.5 ± 12.8 days, and overall, the mortality rate within MV period was 22.6% (72 out of 318). Patient outcomes differed by the presence and absence of PMV, with those that had PMV having a higher mortality rate (38.9% vs. 16.2%, p < 0.001) and longer mean length of ICU stay (19.9 vs. 3.9, p < 0.001). Description of variables used for model development are provided in **Table 1**.

**Table 1.** Description of model co-variables.

| Feature<br>(n = 318) | Presence of<br>Prolonged<br>Mechanical<br>Ventilation<br>(n = 90) | No Presence of<br>Prolonged<br>Mechanical<br>Ventilation<br>(n = 228) | p-value | Total Cohort |
| --- | --- | --- | --- | --- |
| ICU Baseline |  |  |  |  |
| Age | 57.6±16.4 | 60.8±17.1 | 0.125 | 59.9±16.9 |
| Female | 35 (38.9) | 90 (39.5) | 0.923 | 125 (39.3) |
| Admission to medical<br>ICU | 37 (41.1) | 54 (23.7) | 0.002 | 91 (28.6) |
| Primary ICU Admission Diagnosis |  |  |  |  |
| Sepsis/Infection | 7 (7.8) | 14 (6.1) | 0.176 | 21 (6.6) |
| Pulmonary | 10 (11.1) | 13 (5.7) |  | 23 (7.2) |
| Neoplasm | 4 (4.4) | 10 (4.4) |  | 14 (4.4) |
| Gastrointestinal | 6 (6.7) | 16 (7.0) |  | 22 (6.9) |
| Cardiovascular | 11 (12.2) | 64 (28.1) |  | 75 (23.6) |
| Dermatology | 2 (2.2) | 1 (0.4) |  | 3 (0.9) |
| Renal | 1 (1.1) | 3 (1.3) |  | 4 (1.3) |
| Neurology | 14 (15.6) | 34 (14.9) |  | 48 (15.1) |
| Endocrine | 0 (0) | 2 (0.8) |  | 2 (0.6) |
| Trauma | 9 (10.0) | 18 (7.9) |  | 27 (8.5) |
| ICU Type |  |  |  |  |
| Burn | 6 (6.7) | 5 (2.2) | <0.001 | 11 (3.5) |
| Cardiac | 14 (15.6) | 95 (41.7) |  | 109 (34.3) |
| Medical | 37 (41.1) | 54 (23.7) |  | 91 (28.6) |
| Neurosciences | 13 (14.4) | 24 (10.5) |  | 37 (11.6) |
| Surgical | 16 (17.8) | 40 (17.5) |  | 56 (17.6) |
| Mixed | 4 (4.4) | 10 (4.4) |  | 14 (4.4) |
| Mode of Ventilation |  |  |  |  |
| Pressure control | 4 (5.5) | 6 (3.4) | 0.675 | 10 (4.0) |
| Pressure regulated<br>volume control | 61 (83.6) | 134 (80.3) |  | 204 (81.3) |
| SIMV/Pressure<br>regulated volume<br>control | 5 (6.9) | 15 (8.4) |  | 20 (8.0) |
| SIMV/Volume<br>Control | 0 (0) | 3 (1.7) |  | 3 (1.2) |
| Volume Control | 3 (4.1) | 11 (6.2) |  | 14 (5.6) |
| 24 hours after ICU admission |  |  |  |  |
| APACHE II at 24<br>hours | 21.0 ± 5.7 | 20.5 ± 5.3 | 0.443 | 21.0 ± 5.4 |
| SOFA at 24 hours | 10.4 ± 3.9 | 9.8±3.1 | 0.307 | 9.9 ± 3.3 |
| AKI at 24 hours | 34 (37.8) | 26 (11.5) | <0.001 | 90 (28.48) |
| Medication data |  |  |  |  |
| Selected Drugs |  |  |  |  |
| Bumetanide | 5 (5.6) | 4 (1.8) | 0.124 | 9 (2.8) |
| Chlorothiazide | 1 (1.1) | 2 (0.9) | 0.846 | 3 (0.9) |
| Cisatracurium | 4 (4.4) | 4 (1.8) | 0.118 | 8 (2.5) |
| Clonazepam | 0 (0) | 9 (4.0) | 0.065 | 9 (2.8) |
| Dexamethasone | 10 (11.1) | 14 (6.1) | 0.202 | 24 (7.5) |
| Dexmedetomidine | 11 (12.2) | 70 (30.7) | 0.001 | 81 (25.5) |
| Dobutamine | 3 (3.3) | 34 (14.9) | 0.007 | 37 (11.6) |
| Dopamine | 6 (6.7) | 46 (20.2) | 0.003 | 52 (16.4) |
| Ertapenem | 3 (3.3) | 1 (0.4) | 0.070 | 4 (1.3) |
| Epinephrine | 9 (10.0) | 44 (19.3) | 0.045 | 53 (16.7) |
| Hydromorphone | 17 (18.9) | 62 (27.2) | 0.123 | 79 (24.8) |
| Lorazepam | 28 (31.1) | 41 (18.0) | 0.016 | 69 (21.7) |
| Midazolam | 35 (38.9) | 122 (53.5) | 0.019 | 157 (49.4) |
| Milrinone | 1 (1.1) | 25 (11.0) | 0.002 | 26 (8.2) |
| Norepinephrine | 42 (46.7) | 88 (38.6) | 0.233 | 130 (40.9) |
| Phenylephrine | 19 (21.1) | 57 (25.0) | 0.464 | 76 (23.9) |
| Rocuronium | 14 (15.6) | 61 (26.8) | 0.034 | 75 (23.6) |
| Sodium Chloride | 65 (72.2) | 185 (81.1) | 0.081 | 250 (78.6) |
| Vasopressin | 20 (22.2) | 59 (25.9) | 0.497 | 79 (24.8) |
| Neuromuscular blockers | 17 (18.9) | 64 (28.1) | 0.091 | 81 (25.5) |
| Vasopressors | 46 (51.1) | 107 (46.9) | 0.501 | 153 (48.1) |
| Steroids | 25 (27.8) | 72 (31.6) | 0.507 | 97 (30.5) |
| Benzodiazepines | 53 (58.9) | 140 (61.4) | 0.679 | 193 (60.7) |
| Inotropes | 12 (13.3) | 62 (27.2) | 0.008 | 74 (23.3) |
| MRC-ICU at 24 hours of ICU admission | 19.9 ± 10.0 | 21.8 ± 10.7 | 0.153 | 21.3 ± 10.5 |
| MRC-ICU at time of intubation | 17.9 ± 10.7 | 22.6±15.0 | 0.002 | 21.3 ± 14.1 |
| MRC-ICU at 24 hours after intubation | 18.6 ± 10.1 | 17.6±10.7 | 0.464 | 17.9 ± 10.5 |
| MRC-ICU at 24 hours after ICU admission |  |  |  |  |
| <10 | 12 (13.3) | 28 (12.3) | 0.799 | 40 (12.6) |
| ≥10 | 78 (86.7) | 200 (87.7) |  | 278 (87.4) |
| MRC-ICU at time of intubation |  |  |  |  |
| <10 | 21 (23.3) | 52 (22.8) | 0.920 | 73 (23.0) |
| ≥10 | 69 (76.7) | 176 (77.2) |  | 245 (77.0) |
| MRC-ICU at 24 hours after intubation |  |  |  |  |
| <10 | 18 (20.0) | 62 (27.2) | 0.183 | 80 (25.2) |
| ≥10 | 72 (80.0) | 166 (72.8) |  | 238 (74.8) |
| Flowsheet and laboratory values at time of intubation |  |  |  |  |
| Duration of support devices before intubation (HFNC, BiPAP, CPAP) |  |  |  |  |
| 1 day | 10 (58.8) | 21 (70.0) | 0.038 | 31 (66.0) |
| 2 days | 1 (5.9) | 1 (3.3) |  | 2 (4.3) |
| 3 days | 1 (5.9) | 5 (16.7) |  | 6 (12.8) |
| ≥ 4 days | 5 (29.4) | 3 (10.0) |  | 8 (17.0) |
| Elevated pulse n/N (%) | 13/37 (22.2) | 22/77 (28.6) | 0.687 | 35/114 (30.7) |
| HCO<sub>3</sub> < 20 mEq/L n/N (%) | 19/52 (53.3) | 31/110 (18.4) | 0.372 | 50/116 (30.9) |
| PaCO<sub>2</sub> | 36.8 ± 6.3 | 40.4 ± 10.0 | 0.002 | 39.2 ± 9.1 |
| FiO<sub>2</sub> | 55.1 ± 22.3 | 46.7 ± 17.1 | 0.053 | 49.3 ± 19.2 |
| PaO<sub>2</sub>/ FiO<sub>2</sub> median<br>(interquartile range<br>(IQR)) | 320 (216-446) | 307 (237-435) | 0.434 | 308 (234-438) |
| PEEP | 6.6 ± 2.3 | 7.4 ± 3.1 | 0.002 | 6.9 ± 2.6 |
| Minute Ventilation | 6.9 ± 3.4 | 7.0 ± 3.9 | 0.779 | 7.0 ± 3.7 |
| PaCO<sub>2</sub> * Minute<br>Ventilation | 255.2 ± 131.4 | 285.1 ± 192.0 | 0.346 | 272.3 ± 168.6 |
| Fluid overload n/N<br>(%) | 17/90 (57.8) | 23/227 (19.3) | 0.054 | 40/317 (12.6) |
| *Data are presented as n (%) or mean ± std unless otherwise stated.*<br>*SOFA: sequential organ failure assessment, APACHE II: Acute Physiology and Chronic Health Evaluation; ICU: intensive care unit; PEEP: positive end expiratory pressure; SIMV: synchronized intermittent mandatory ventilation* |  |  |  |  |

When predicting PMV, statistical and machine learning models were developed, and model performance is summarized in **Table 2**. Using logistic regression, a base model was developed and demonstrated an AUROC of 0.67 (0.39 – 0.94) with an accuracy of 0.78 (0.60 – 0.90). A series of candidate models were then developed using MRC-ICU and severity of illness scores. The nested model using the MRC-ICU at 24 hours was superior to the others and following variable selection, AUROC was improved to 0.73 (0.45 – 0.99) and accuracy of 0.85 (0.68 – 0.95) with a sensitivity of 0.63 and specificity of 0.91, respectively, when Youden’s threshold was applied. No evidence of collinearity was found among the predictors. Results for the full regression model are provided in **Supplemental Content – Table 1**. The mediation analysis showed duration of support devices and HCO_3_ are mediators between the outcome and MRC-ICU (p-value = 0.016 and <0.001, respectively). After reviewing the effect of the three mediators, the final selected model in predicting PMV was developed (see Supplemental Content - Table 2).

**Table 2.**
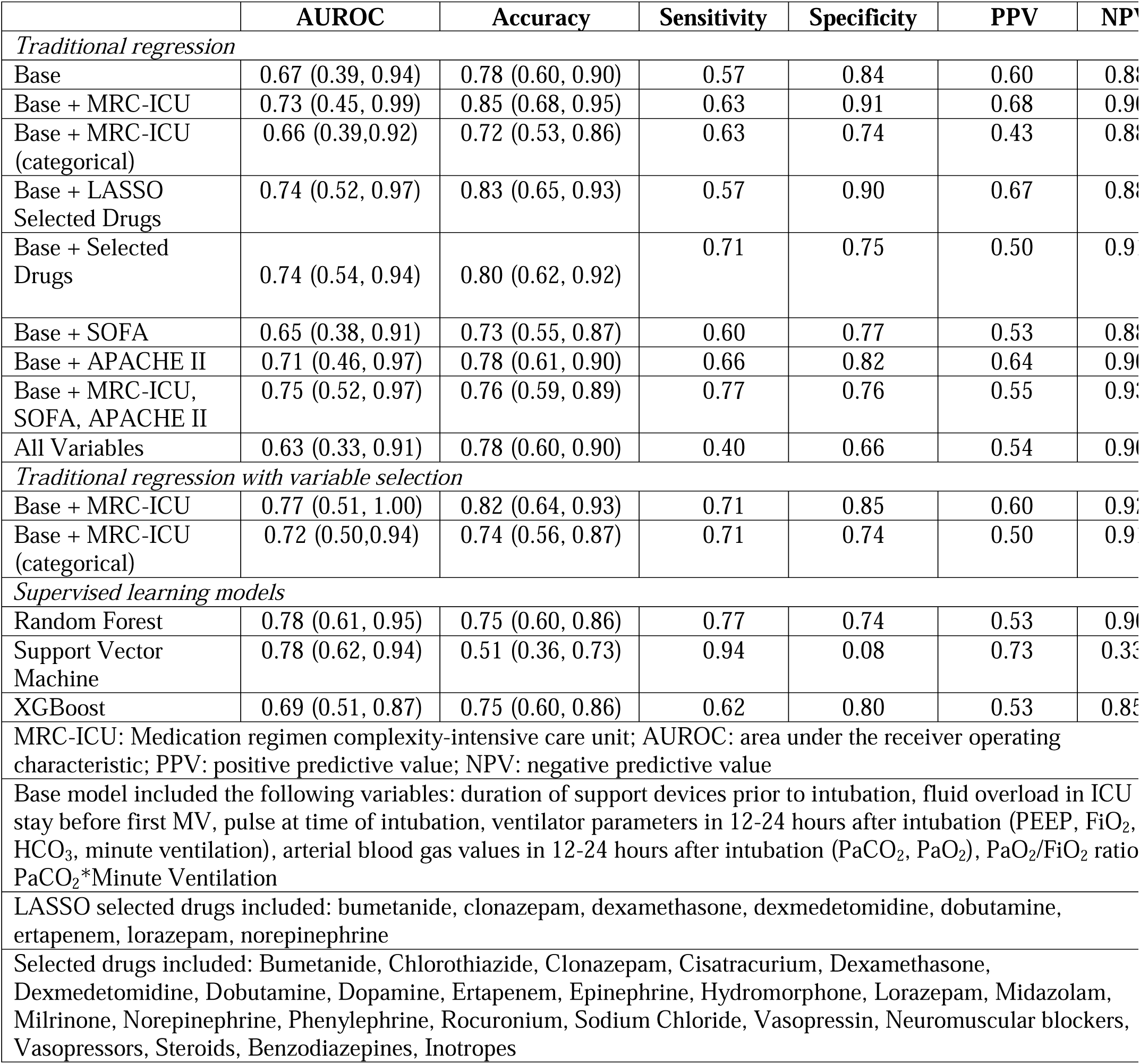
Performance of prolonged duration of mechanical ventilation prediction models.

Among the machine learning models, Random Forest and SVM shared the highest AUROC (0.78) compared to XGBoost (0.69); however, the SVM model was limited by a relatively low accuracy (0.51), potentially secondary to imbalanced data due to the lower overall rate of PMV. To test this, we used smote function in R to oversample PMV and create a balanced dataset. With additional observations of PMV, the accuracy increased to 0.70, which suggested that the imbalanced outcome is one of the reasons leading to the low accuracy of SVM model. Model performance is summarized in **Table 2**. AUROC graphs are plotted in Figure 2. Finally, feature importance graphs were plotted for Random Forest (see Figure 3 **), SVM** (see **Supplemental Content -** Figure 2), and XGBoost (see **Supplemental Content -** Figure 3). The top five features for each analysis were plotted by count. For Random Forest, the top five included duration of support devices prior to intubation, SOFA score at 24 hours, PaO_2_/FiO_2_ ratio, MRC-ICU score at the time of intubation, and PaCO_2_, with MRC-ICU at 24 hours and a product of minute ventilation times PaCO_2_ scoring in the top ten. For SVM, the top five most important features (in descending order) were duration of support devices prior to intubation, PaCO_2_, elevated pulse, midazolam, and dexmedetomidine, though again MRC-ICU at 24 hours and the time of intubation also were in the top ten. For XGBoost, the top 5 features (in descending order) were similar: MRC-ICU at intubation, minute ventilation, PaCO_2_, a product of minute ventilation times paCO_2,_ and SOFA, with again MRC-ICU at 24 hours and 24 hours after intubation scoring in the top ten. These variables had overlap with the regression model, with arterial blood gas values, ventilator settings, and MRC-ICU at the time of intubation being included.

**Figure 2.**
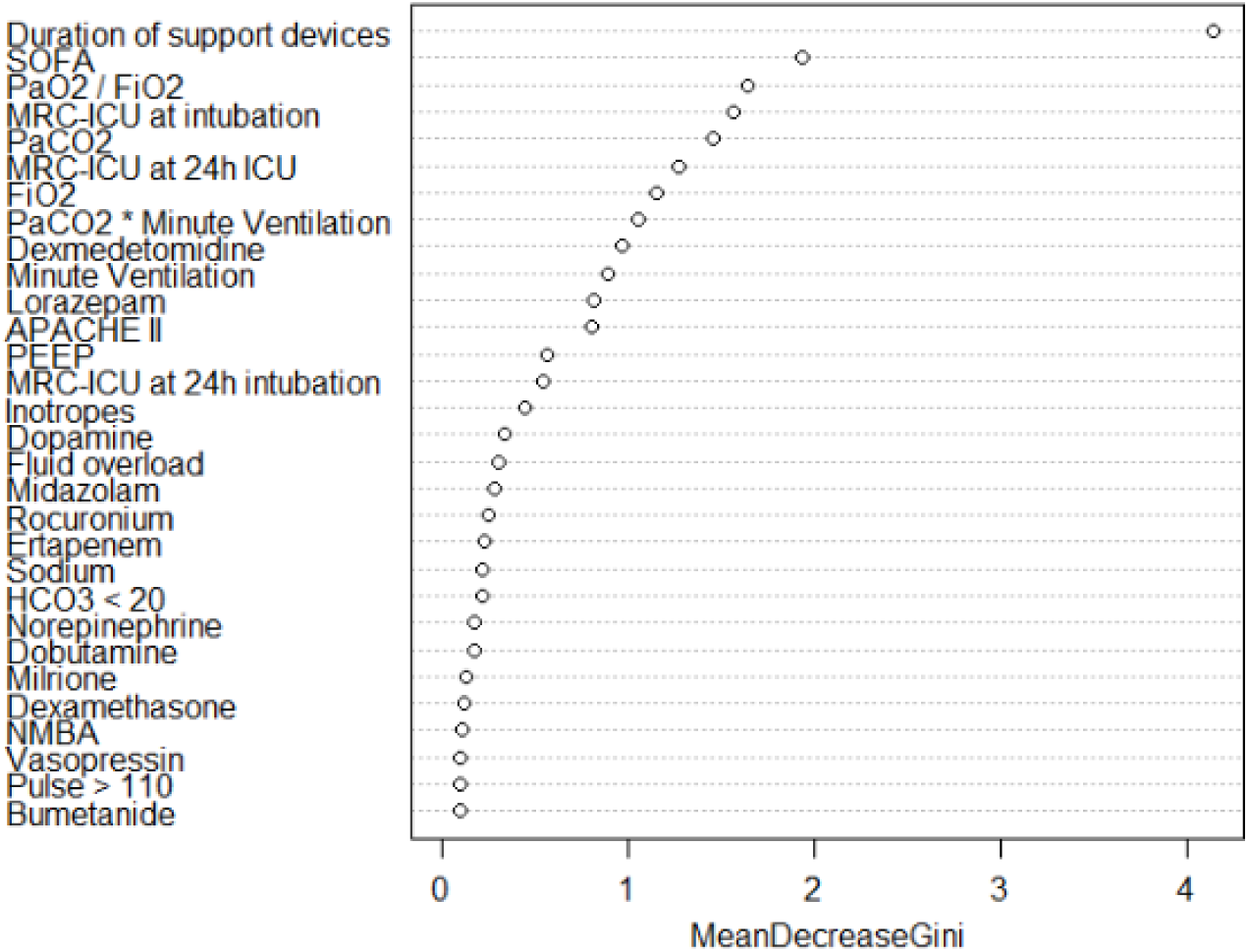
Feature importance graph using Random Forest.

## Discussion

In the first machine learning analysis for predicting PMV risk in critically ill patients to include detailed medication data, both logistic regression and machine learning analyses demonstrated similar predictive power to identify patients at risk of PMV at the end of the first 24 hours of their ICU stay. Compared to SVM and XGBoost, Random Forest displayed the more robust predictive capabilities, balancing AUROC with reasonable accuracy and high NPV. The machine learning analyses revealed differences with regard to the feature importance variables compared to logistic regression, with the importance of medication-related variables as top predictors repeatedly observed. This finding is particularly salient because while certain medications are known to prolong duration of mechanical ventilation, they are not frequently captured in other analyses that have aimed to predict PMV.

In particular, the logistic regression model with MRC-ICU and Random Forest model achieved reasonable AUROCs, and negative predictive values were high for all models developed. These results are in line with previous evaluations PMV (see **Table 3**), notably with relatively high NPV. (10,11,26–28,31,36,37) Several machine learning analyses of PMV have been conducted in narrow populations (i.e., traumatic brain injury, congestive heart failure, after coronary artery bypass grafting) or used to predict mortality, whereas the present study evaluated a diverse scope of critically ill patients across a variety of ICU settings, which may support external generalizability. (38–41) Moreover, some studies (see **Table 3**) have included elements of medication therapy in their modeling (e.g., duration or dependency on inotropes), but to date, none have taken into account the entire medication administration record or attempted to quantify the additive effects of various medications, as the MRC-ICU has been validated to do. (11,16,29,42) The studies outlined in **Table 3** incorporated traditional statistical modeling to evaluate predictors of PMV; however, the majority of studies that evaluated medication use found that variable nonsignificant in multivariate analysis. (28,31,33,37) Notably, one study evaluating use of common sedatives, vasopressors, steroids, and neuromuscular blocking agents found the duration of vasopressor support to be an independent predictor of PMV. (11)

**Table 3.**
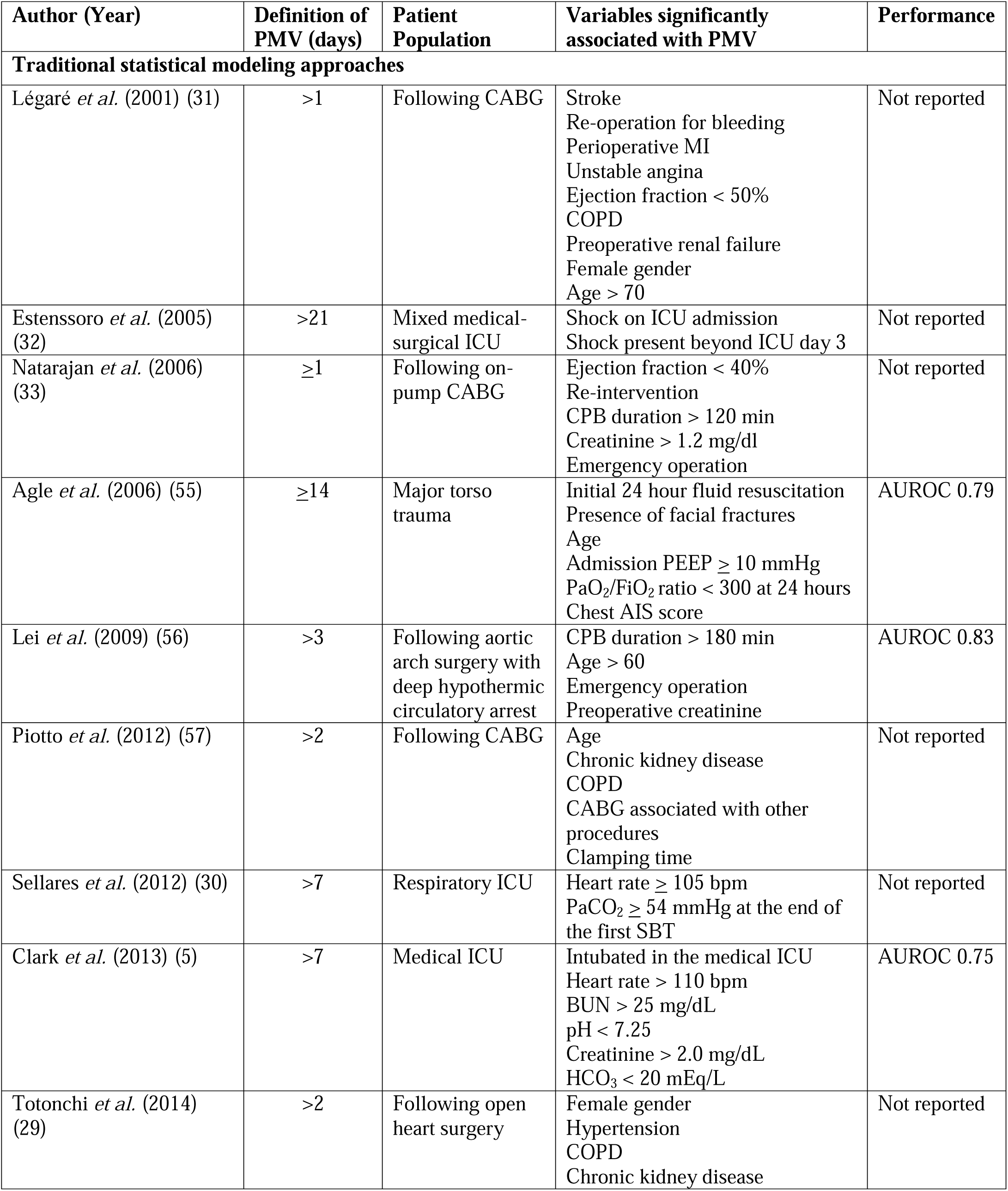
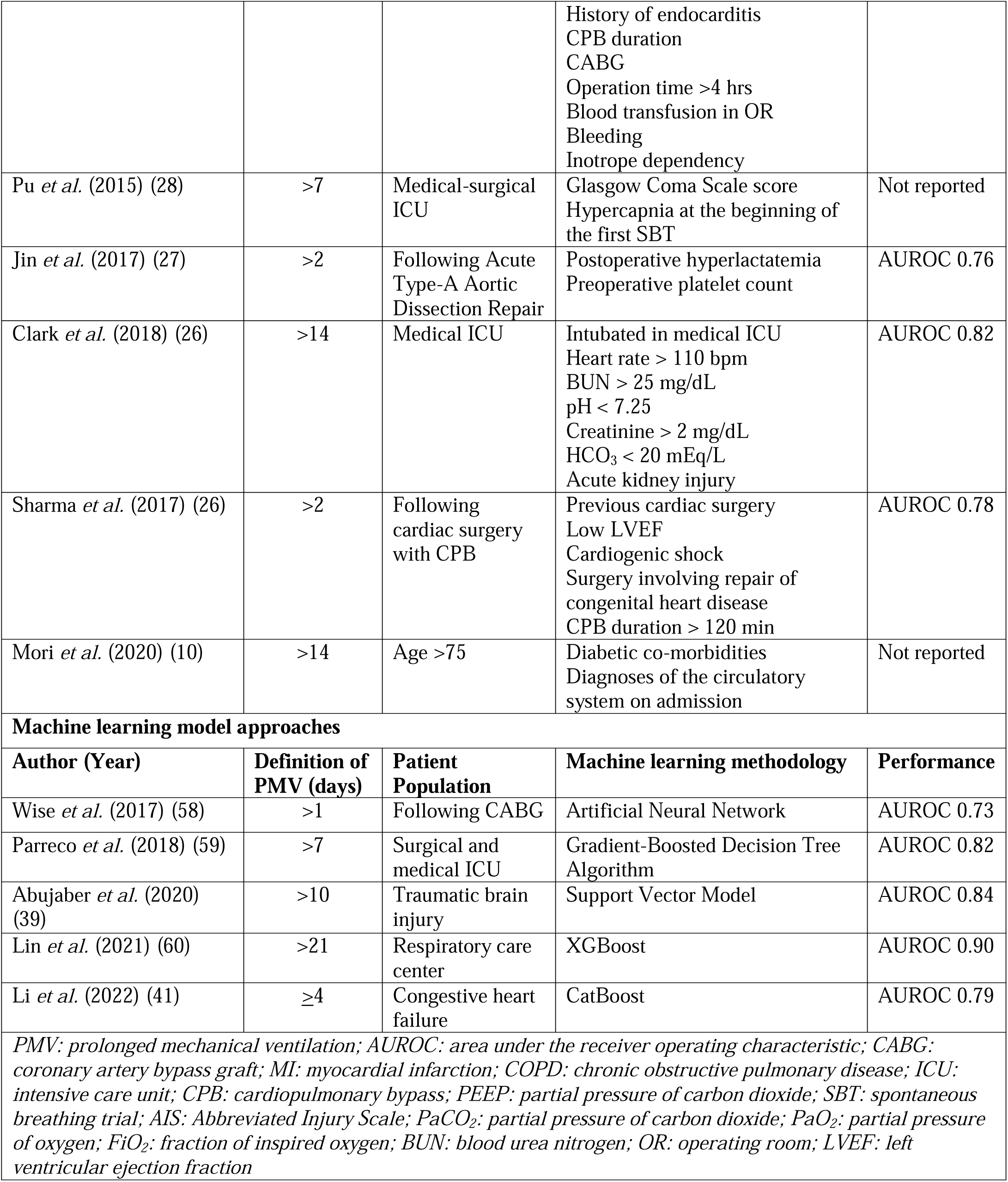
Summary of studies evaluating prolonged mechanical ventilation.

| Author (Year) | Definition of PMV (days) | Patient Population | Variables significantly associated with PMV | Performance |
| --- | --- | --- | --- | --- |
| **Traditional statistical modeling approaches** |  |  |  |  |
| Légaré *et al.* (2001) (31) | >1 | Following CABG | Stroke<br>Re-operation for bleeding<br>Perioperative MI<br>Unstable angina<br>Ejection fraction < 50%<br>COPD<br>Preoperative renal failure<br>Female gender<br>Age > 70 | Not reported |
| Estenssoro *et al.* (2005) (32) | >21 | Mixed medical-surgical ICU | Shock on ICU admission<br>Shock present beyond ICU day 3 | Not reported |
| Natarajan *et al.* (2006) (33) | $\geq 1$ | Following on-pump CABG | Ejection fraction < 40%<br>Re-intervention<br>CPB duration > 120 min<br>Creatinine > 1.2 mg/dl<br>Emergency operation | Not reported |
| Agle *et al.* (2006) (55) | $\geq 14$ | Major torso trauma | Initial 24 hour fluid resuscitation<br>Presence of facial fractures<br>Age<br>Admission PEEP $\geq 10$ mmHg<br>PaO<sub>2</sub>/FiO<sub>2</sub> ratio < 300 at 24 hours<br>Chest AIS score | AUROC 0.79 |
| Lei *et al.* (2009) (56) | >3 | Following aortic arch surgery with deep hypothermic circulatory arrest | CPB duration > 180 min<br>Age > 60<br>Emergency operation<br>Preoperative creatinine | AUROC 0.83 |
| Piotto *et al.* (2012) (57) | >2 | Following CABG | Age<br>Chronic kidney disease<br>COPD<br>CABG associated with other procedures<br>Clamping time | Not reported |
| Sellares *et al.* (2012) (30) | >7 | Respiratory ICU | Heart rate $\geq 105$ bpm<br>PaCO<sub>2</sub> $\geq 54$ mmHg at the end of the first SBT | Not reported |
| Clark *et al.* (2013) (5) | >7 | Medical ICU | Intubated in the medical ICU<br>Heart rate > 110 bpm<br>BUN > 25 mg/dL<br>pH < 7.25<br>Creatinine > 2.0 mg/dL<br>HCO<sub>3</sub> < 20 mEq/L | AUROC 0.75 |
| Totonchi *et al.* (2014) (29) | >2 | Following open heart surgery | Female gender<br>Hypertension<br>COPD<br>Chronic kidney disease | Not reported |
|  |  |  | History of endocarditis<br>CPB duration<br>CABG<br>Operation time >4 hrs<br>Blood transfusion in OR<br>Bleeding<br>Inotrope dependency |  |
| Pu *et al.* (2015) (28) | >7 | Medical-surgical ICU | Glasgow Coma Scale score<br>Hypercapnia at the beginning of the first SBT | Not reported |
| Jin *et al.* (2017) (27) | >2 | Following Acute Type-A Aortic Dissection Repair | Postoperative hyperlactatemia<br>Preoperative platelet count | AUROC 0.76 |
| Clark *et al.* (2018) (26) | >14 | Medical ICU | Intubated in medical ICU<br>Heart rate > 110 bpm<br>BUN > 25 mg/dL<br>pH < 7.25<br>Creatinine > 2 mg/dL<br>HCO<sub>3</sub> < 20 mEq/L<br>Acute kidney injury | AUROC 0.82 |
| Sharma *et al.* (2017) (26) | >2 | Following cardiac surgery with CPB | Previous cardiac surgery<br>Low LVEF<br>Cardiogenic shock<br>Surgery involving repair of congenital heart disease<br>CPB duration > 120 min | AUROC 0.78 |
| Mori *et al.* (2020) (10) | >14 | Age >75 | Diabetic co-morbidities<br>Diagnoses of the circulatory system on admission | Not reported |
| **Machine learning model approaches** |  |  |  |  |
| **Author (Year)** | **Definition of PMV (days)** | **Patient Population** | **Machine learning methodology** | **Performance** |
| Wise *et al.* (2017) (58) | >1 | Following CABG | Artificial Neural Network | AUROC 0.73 |
| Parreco *et al.* (2018) (59) | >7 | Surgical and medical ICU | Gradient-Boosted Decision Tree Algorithm | AUROC 0.82 |
| Abujaber *et al.* (2020) (39) | >10 | Traumatic brain injury | Support Vector Model | AUROC 0.84 |
| Lin *et al.* (2021) (60) | >21 | Respiratory care center | XGBoost | AUROC 0.90 |
| Li *et al.* (2022) (41) | ≥4 | Congestive heart failure | CatBoost | AUROC 0.79 |
| *PMV: prolonged mechanical ventilation; AUROC: area under the receiver operating characteristic; CABG: coronary artery bypass graft; MI: myocardial infarction; COPD: chronic obstructive pulmonary disease; ICU: intensive care unit; CPB: cardiopulmonary bypass; PEEP: partial pressure of carbon dioxide; SBT: spontaneous breathing trial; AIS: Abbreviated Injury Scale; PaCO<sub>2</sub>: partial pressure of carbon dioxide; PaO<sub>2</sub>: partial pressure of oxygen; FiO<sub>2</sub>: fraction of inspired oxygen; BUN: blood urea nitrogen; OR: operating room; LVEF: left ventricular ejection fraction* |  |  |  |  |

In contrast to the decision to extubate (i.e., a de-escalation of care in a stable patient that can be delayed in situations of high uncertainty), the decision to intubate is an escalation of care often performed in a generally time-sensitive scenario for an unstable patient. Thus, while the rapid shallow breathing index to predict extubation success has a PPV of approximately 78%, it is still a widely used tool. (43) Given the present findings, the ability to predict a high risk of PMV with certainty is limited, and thus as a guide to decisions like early tracheostomy it is likely not viable; however, high NPV may be useful in guiding medication-related decisions (e.g., the decision to use light sedation or be aggressive with diuresis knowing that extubation within 5 days is likely) because it serves as a screening test for PMV. The clinical utility of a model with high NPV is increased clinician confidence that despite the present requirement to intubate the patient, this need is transient. As such, it may direct goals of care discussions and more importantly may guide a more aggressive weaning strategy, both regarding ventilator settings and medication therapy. Indeed, even hours on non-optimized ventilator settings are associated with worse outcomes, including heightened mortality risk. (44–46)

Ventilator settings and medication therapy do not occur in silos but are highly intertwined: liberalizing ventilator settings can reduce the need for sedation (which is known to prolong mechanical ventilation and reduce extubation success). (2,47,48) As opposed to “settling in” for an intubation that is expected to last for an extended period, such a model may guard against clinical inertia towards more active interventions to get the patient successfully extubated (e.g., early mobility, avoidance of benzodiazepines). (49,50) Despite knowing the importance of light sedation and early mobility, translating this knowledge into action has remained a challenge, but quantitative risk prediction data indicating a short intubation period may guard against this inertia. (50) Beyond the intertwined role of drug and device, medications are a unique element to predictive models, because they represent both independent risk factors for PMV but also potentially modifiable ones. It has been previously proposed that medications represent intervenable events in the ICU. Intervenable events have three characteristics: without action by a clinician, they are associated with *poor* outcomes; however, they are both *preventable* and *predictable*. (51) For example, an idiosyncratic, allergic reaction to the antibiotic cefepime would not be intervenable; however, rapid recognition of sepsis that reduces time to cefepime is intervenable. It is well-established that medications play an integral role in the management and outcomes of mechanically ventilated patients, but in contrast to initial severity of illness indicators (repeatedly demonstrated to both predict and cause PMV), they are frequently modifiable, with appropriate clinical decision support. (14,52,53) For example, diuretics and aggressive volume management are associated with reduced time on the ventilator. (54) Interestingly, the MRC-ICU at the time intubation had a negative association with PMV while the MRC-ICU at 24 hours had a positive association with PMV, indicating the possibility that high complexity may at times be a protective factor. As such, models that incorporate medications present a unique opportunity to guide clinical decision-making that reduces duration of mechanical ventilation.

Limitations of this study include sample size, which though reasonable within many critical care evaluations is comparatively small for machine learning applications. While validation and training sets were used, this evaluation lacks external validation in a separate dataset, and the results do not address the means by which to operationalize such a prediction metric at the bedside. Finally, while the presence or absence of certain medications appeared to play a role in PMV, timing and dosing of these medications (also known to be important for intubated patients) were not evaluated at this juncture.

## Conclusion

PMV is a potentially intervenable event in the ICU population with the appropriate prediction modeling that can guide timely intervention. The incorporation of medication-related variables and exploration modeling techniques is an important step towards the prevention of this ICU complication.

## Data Availability

All data produced in the present study are available upon reasonable request to the authors.

## Acknowledgements

Data acquisition were supported by NC TraCS, funded by Grant Number UL1TR002489 from the National Center for Advancing Translations Sciences at the National Institutes of Health, and Data Analytics at the University of North Carolina Medical Center Department of Pharmacy. American Society of Health-System Pharmacists Innovations in Technology Grant supported this work.

## References

1. Cawley MJ. Mechanical ventilation: introduction for the pharmacy practitioner. J Pharm Pract 2011;24(1):7–16.

2. Murray B, Sikora A, Mock JR, Devlin T, Keats K, Powell R, et al. Reverse Triggering: An Introduction to Diagnosis, Management, and Pharmacologic Implications. Front Pharmacol 2022;13:879011.

3. Chant C, Dewhurst NF, Friedrich JO. Do we need a pharmacist in the ICU? Intensive Care Med 2015;41(7):1314–1320.

4. Marshall J, Finn CA, Theodore AC. Impact of a clinical pharmacist-enforced intensive care unit sedation protocol on duration of mechanical ventilation and hospital stay. Crit Care Med 2008;36(2):427–433.

5. Clark PA, Lettieri CJ. Clinical model for predicting prolonged mechanical ventilation. J Crit Care 2013;28(5):880 e881-887.

6. Cawley MJ. Mechanical ventilation: a tutorial for pharmacists. Pharmacotherapy 2007;27(2):250–266.

7. Cawley MJ. Advanced Modes of Mechanical Ventilation: Introduction for the Critical Care Pharmacist. J Pharm Pract 2019;32(2):186–198.

8. Newsome AS, Chastain, D. B., Watkins, P., & Hawkins, W. A.. Complications and Pharmacologic Interventions of Invasive Positive Pressure Ventilation During Critical Illness.. Journal of Pharmacy Technology 2018;34(4):153–170.

9. Ghauri SK, Javaeed A, Mustafa KJ, Khan AS. Predictors of prolonged mechanical ventilation in patients admitted to intensive care units: A systematic review. Int J Health Sci (Qassim) 2019;13(6):31–38.

10. Mori H, Yamasaki K, Itoh T, Saishoji Y, Torisu Y, Mori T, et al. Predictors of prolonged mechanical ventilation identified at an emergency visit for elderly people: A retrospective cohort study. Medicine (Baltimore) 2020;99(49):e23472.

11. Muzaffar SN, Gurjar M, Baronia AK, Azim A, Mishra P, Poddar B, et al. Predictors and pattern of weaning and long-term outcome of patients with prolonged mechanical ventilation at an acute intensive care unit in North India. Rev Bras Ter Intensiva 2017;29(1):23–33.

12. Uijtendaal EV, van Harssel LL, Hugenholtz GW, Kuck EM, Zwart-van Rijkom JE, Cremer OL, et al. Analysis of potential drug-drug interactions in medical intensive care unit patients. Pharmacotherapy 2014;34(3):213–219.

13. Newsome A, Smith, SE, Jones, TW, Taylor, A, Van Berkel, MA, Rabinovich, M. . A survey of critical care pharmacists to patient ratios and practice characteristics in intensive care units. J Am Coll Clin Pharm 2020;3:68-74,.

14. Stollings JL, Foss JJ, Ely EW, Ambrose AM, Rice TW, Girard TD, et al. Pharmacist leadership in ICU quality improvement: coordinating spontaneous awakening and breathing trials. Ann Pharmacother 2015;49(8):883–891.

15. Leguelinel-Blache G, Nguyen TL, Louart B, Poujol H, Lavigne JP, Roberts JA, et al. Impact of Quality Bundle Enforcement by a Critical Care Pharmacist on Patient Outcome and Costs. Crit Care Med 2018;46(2):199–207.

16. Sikora A, Ayyala D, Rech MA, Blackwell SB, Campbell J, Caylor MM, et al. Impact of Pharmacists to Improve Patient Care in the Critically Ill: A Large Multicenter Analysis Using Meaningful Metrics With the Medication Regimen Complexity-ICU (MRC-ICU) Score. Crit Care Med 2022;50(9):1318–1328.

17. Newsome AS, Smith SE, Olney WJ, Jones TW. Multicenter validation of a novel medication-regimen complexity scoring tool. Am J Health Syst Pharm 2020;77(6):474–478.

18. Newsome AS, Anderson D, Gwynn ME, Waller JL. Characterization of changes in medication complexity using a modified scoring tool. Am J Health Syst Pharm 2019;76(Supplement_4):S92-s95.

19. Gwynn ME, Poisson MO, Waller JL, Newsome AS. Development and validation of a medication regimen complexity scoring tool for critically ill patients. Am J Health Syst Pharm 2019;76(Supplement_2):S34-s40.

20. Al-Mamun MA, Brothers T, Newsome AS. Development of Machine Learning Models to Validate a Medication Regimen Complexity Scoring Tool for Critically Ill Patients. Ann Pharmacother 2021;55(4):421–429.

21. Smith SE, Shelley R, Sikora A. Medication regimen complexity vs patient acuity for predicting critical care pharmacist interventions. Am J Health Syst Pharm 2022;79(8):651–655.

22. Webb AJ, Rowe S, Newsome AS. A descriptive report of the rapid implementation of automated MRC-ICU calculations in the EMR of an academic medical center. Am J Health Syst Pharm 2022;79(12):979–983.

23. Newsome AS, Smith SE, Olney WJ, Jones TW, Forehand CC, Jun AH, et al. Medication regimen complexity is associated with pharmacist interventions and drug-drug interactions: A use of the novel MRC-ICU scoring tool. JACCP: JOURNAL OF THE AMERICAN COLLEGE OF CLINICAL PHARMACY 2020;3(1):47–56.

24. Olney WJ, Chase AM, Hannah SA, Smith SE, Newsome AS. Medication Regimen Complexity Score as an Indicator of Fluid Balance in Critically Ill Patients. J Pharm Pract 2021:897190021999792.

25. Murray B, Zhao B, Kong Y, Shen Y, Sikora A. 935: PREDICTING DURATION OF MECHANICAL VENTILATION WITH MEDICATION REGIMEN COMPLEXITY VARIABLES. Critical Care Medicine 2023;51(1):460.

26. Sharma V, Rao V, Manlhiot C, Boruvka A, Fremes S, Wasowicz M. A derived and validated score to predict prolonged mechanical ventilation in patients undergoing cardiac surgery. J Thorac Cardiovasc Surg 2017;153(1):108–115.

27. Jin M, Ma WG, Liu S, Zhu J, Sun L, Lu J, et al. Predictors of Prolonged Mechanical Ventilation in Adults After Acute Type-A Aortic Dissection Repair. J Cardiothorac Vasc Anesth 2017;31(5):1580–1587.

28. Pu L, Zhu B, Jiang L, Du B, Zhu X, Li A, et al. Weaning critically ill patients from mechanical ventilation: A prospective cohort study. J Crit Care 2015;30(4):862 e867-813.

29. Totonchi Z, Baazm F, Chitsazan M, Seifi S, Chitsazan M. Predictors of prolonged mechanical ventilation after open heart surgery. J Cardiovasc Thorac Res 2014;6(4):211–216.

30. Sellares J, Ferrer M, Cano E, Loureiro H, Valencia M, Torres A. Predictors of prolonged weaning and survival during ventilator weaning in a respiratory ICU. Intensive Care Med 2011;37(5):775–784.

31. Legare JF, Hirsch GM, Buth KJ, MacDougall C, Sullivan JA. Preoperative prediction of prolonged mechanical ventilation following coronary artery bypass grafting. Eur J Cardiothorac Surg 2001;20(5):930–936.

32. Estenssoro E, Gonzalez F, Laffaire E, Canales H, Saenz G, Reina R, et al. Shock on admission day is the best predictor of prolonged mechanical ventilation in the ICU. Chest 2005;127(2):598–603.

33. Natarajan K, Patil S, Lesley N, Ninan B. Predictors of prolonged mechanical ventilation after on-pump coronary artery bypass grafting. Ann Card Anaesth 2006;9(1):31–36.

34. White IR RP, Wood AM. Multiple imputation using chained, 10.1002/sim.4067eiagfpSMd.

35. Rubin DB. Multiple imputation for nonresponse in surveys. Hoboken, N.J. ;: Wiley-Interscience; 2004.

36. Clark PA, Inocencio RC, Lettieri CJ. I-TRACH: Validating A Tool for Predicting Prolonged Mechanical Ventilation. J Intensive Care Med 2018;33(10):567–573.

37. Cottereau G, Dres M, Avenel A, Fichet J, Jacobs FM, Prat D, et al. Handgrip Strength Predicts Difficult Weaning But Not Extubation Failure in Mechanically Ventilated Subjects. Respir Care 2015;60(8):1097–1104.

38. Abujaber A, Fadlalla A, Gammoh D, Abdelrahman H, Mollazehi M, El-Menyar A. Prediction of in-hospital mortality in patients on mechanical ventilation post traumatic brain injury: machine learning approach. BMC Med Inform Decis Mak 2020;20(1):336.

39. Abujaber A, Fadlalla A, Gammoh D, Abdelrahman H, Mollazehi M, El-Menyar A. Using trauma registry data to predict prolonged mechanical ventilation in patients with traumatic brain injury: Machine learning approach. PLoS One 2020;15(7):e0235231.

40. George N, Moseley E, Eber R, Siu J, Samuel M, Yam J, et al. Deep learning to predict long-term mortality in patients requiring 7 days of mechanical ventilation. PLoS One 2021;16(6):e0253443.

41. Li L, Tu B, Xiong Y, Hu Z, Zhang Z, Liu S, et al. Machine Learning-Based Model for Predicting Prolonged Mechanical Ventilation in Patients with Congestive Heart Failure. Cardiovasc Drugs Ther 2022.

42. Olney WJ, Chase AM, Hannah SA, Smith SE, Newsome AS. Medication Regimen Complexity Score as an Indicator of Fluid Balance in Critically Ill Patients. J Pharm Pract 2022;35(4):573–579.

43. Yang KL, Tobin MJ. A prospective study of indexes predicting the outcome of trials of weaning from mechanical ventilation. N Engl J Med 1991;324(21):1445–1450.

44. Needham DM, Yang T, Dinglas VD, Mendez-Tellez PA, Shanholtz C, Sevransky JE, et al. Timing of low tidal volume ventilation and intensive care unit mortality in acute respiratory distress syndrome. A prospective cohort study. Am J Respir Crit Care Med 2015;191(2):177–185.

45. Page D, Ablordeppey E, Wessman BT, Mohr NM, Trzeciak S, Kollef MH, et al. Emergency department hyperoxia is associated with increased mortality in mechanically ventilated patients: a cohort study. Crit Care 2018;22(1):9.

46. Gajic O, Frutos-Vivar F, Esteban A, Hubmayr RD, Anzueto A. Ventilator settings as a risk factor for acute respiratory distress syndrome in mechanically ventilated patients. Intensive Care Med 2005;31(7):922–926.

47. Newsome AS, Chastain DB, Watkins P, Hawkins WA. Complications and Pharmacologic Interventions of Invasive Positive Pressure Ventilation During Critical Illness. J Pharm Technol 2018;34(4):153–170.

48. Devlin JW, Skrobik Y, Gelinas C, Needham DM, Slooter AJC, Pandharipande PP, et al. Clinical Practice Guidelines for the Prevention and Management of Pain, Agitation/Sedation, Delirium, Immobility, and Sleep Disruption in Adult Patients in the ICU. Crit Care Med 2018;46(9):e825–e873.

49. Marra A, Ely EW, Pandharipande PP, Patel MB. The ABCDEF Bundle in Critical Care. Crit Care Clin 2017;33(2):225–243.

50. Ely EW. Every deep-drawn breath : a critical care doctor on healing, recovery, and transforming medicine in the ICU. In: First Scribner hardcover edition. ed. New York: Scribner,; 2021.

51. Sikora A. Critical Care Pharmacists: A Focus on Horizons. Critical Care Clinics 2023;[In press].

52. Lightfoot M, Sanders A, Burke C, Patton J. Clinical Pharmacist Impact on Intensive Care Unit Delirium: Intervention and Monitoring. Hosp Pharm 2019;54(3):180–185.

53. Zhang Z, Chen K, Ni H, Zhang X, Fan H. Sedation of mechanically ventilated adults in intensive care unit: a network meta-analysis. Sci Rep 2017;7:44979.

54. Bissell BD, Laine ME, Thompson Bastin ML, Flannery AH, Kelly A, Riser J, et al. Impact of protocolized diuresis for de-resuscitation in the intensive care unit. Crit Care 2020;24(1):70.

55. Agle SC, Kao LS, Moore FA, Gonzalez EA, Vercruysse GA, Todd SR. Early predictors of prolonged mechanical ventilation in major torso trauma patients who require resuscitation. Am J Surg 2006;192(6):822–827.

56. Lei Q, Chen L, Zhang Y, Fang N, Cheng W, Li L. Predictors of prolonged mechanical ventilation after aortic arch surgery with deep hypothermic circulatory arrest plus antegrade selective cerebral perfusion. J Cardiothorac Vasc Anesth 2009;23(4):495–500.

57. Piotto RF, Ferreira FB, Colosimo FC, Silva GS, Sousa AG, Braile DM. Independent predictors of prolonged mechanical ventilation after coronary artery bypass surgery. Rev Bras Cir Cardiovasc 2012;27(4):520–528.

58. Wise ES, Stonko DP, Glaser ZA, Garcia KL, Huang JJ, Kim JS, et al. Prediction of Prolonged Ventilation after Coronary Artery Bypass Grafting: Data from an Artificial Neural Network. Heart Surg Forum 2017;20(1):E007–e014.

59. Parreco J, Hidalgo A, Parks JJ, Kozol R, Rattan R. Using artificial intelligence to predict prolonged mechanical ventilation and tracheostomy placement. J Surg Res 2018;228:179–187.

60. Lin MY, Li CC, Lin PH, Wang JL, Chan MC, Wu CL, et al. Explainable Machine Learning to Predict Successful Weaning Among Patients Requiring Prolonged Mechanical Ventilation: A Retrospective Cohort Study in Central Taiwan. Front Med (Lausanne) 2021;8:663739.

