## Supplementary material for "Machine learning based prediction of prolonged duration of mechanical ventilation incorporating medication data": Online Supplement

**Table E1.** Univariate and multivariate analysis for prolonged mechanical ventilation

**Table E2.** Logistic regression for final model to predict prolonged mechanical ventilation

**Figure E3.** The flowchart of model development.

**Figure E4.** Feature importance graph using Support Vector Machine

**Figure E5.** Feature importance graph using XGBoost

**Table E1.** Univariate and multivariate analysis for prolonged mechanical ventilation

|  | **Univariate** | | | **Multivariate** | | |
| --- | --- | --- | --- | --- | --- | --- |
|  | **Odds Ratio** | **95% CI** | **p-value** | **Odds Ratio** | **95% CI** | **p-value** |
| **ICU Baseline** |  |  |  |  |  |  |
| Age | 0.98 | (0.97, 1.00) | 0.12 | 0.87 | (0.52, 1.44) | 0.58 |
| Female | 0.97 | (0.59, 1.61) | 0.92 | 0.87 | (0.39, 1.91) | 0.72 |
| Admission to medical ICU | 2.24 | (1.34, 3.78) | <0.01 | 1.44 | (0.44, 4.70) | 0.55 |
| *Primary ICU Admission Diagnosis* | | | | | | |
| Sepsis/Infection | 1.28 | (0.5, 3.31) | 0.59 | 0.34 | (0.02, 6.14) | 0.49 |
| Pulmonary | 2.06 | (0.87, 4.9) | 0.09 | 3.68 | (0.64, 21.10) | 0.15 |
| Neoplasm | 1.01 | (0.31, 3.32) | 0.98 | 0.27 | (0.01, 7.92) | 0.46 |
| Gastrointestinal | 0.94 | (0.36, 2.5) | 0.91 | 0.50 | (0.07, 3.75) | 0.51 |
| Cardiovascular | 0.35 | (0.18, 0.71) | <0.01 | 0.56 | (0.09, 3.49) | 0.54 |
| Dermatology | 5.15 | (0.46, 57.62) | 0.18 | 11.52 | (0.54, 246.9) | 0.12 |
| Renal | 0.84 | (0.09, 8.21) | 0.88 | 1.21 | (0.03, 43.37) | 0.92 |
| Neurology | 1.05 | (0.53, 2.07) | 0.88 | 0.37 | (0.07, 1.86) | 0.24 |
| Endocrine | 1.19E-06 | (0, Inf) | 0.98 | 1.40E-07 | (0, Inf) | 0.99 |
| Trauma | 1.29 | (0.56, 3.00) | 0.54 | 0.73 | (0.12, 4.56) | 0.74 |
| **24 hours after ICU admission** | | | | | | |
| APACHE II at 24 hours | 1.01 | (0.97, 1.07) | 0.44 | 1.26 | (0.68, 2.34) | 0.47 |
| SOFA at 24 hours | 1.05 | (0.96, 1.16) | 0.25 | 1.17 | (0.45, 3.01) | 0.76 |
| Fluid overload at MV start /N (%) | 2.06 | (1.04, 4.08) | 0.03 | 0.33 | (0.09, 1.31) | 0.12 |
| AKI at 24 hours | 4.67 | (2.59, 8.43) | <0.01 | 9.41 | (2.98, 29.71) | <0.01 |
| **Medication data** | | | | | | |
| *Selected Drugs* | | | | | | |
| Bumetanide | 3.29 | (0.86, 12.56) | 0.08 | 5.03 | (0.37, 67.79) | 0.23 |
| Chlorothiazide | 1.27 | (0.11, 14.18) | 0.42 | 3.60 | (0.04, 319.4) | 0.58 |
| Clonazepam | 1.55E-07 | (0, Inf) | 0.98 | 6.69E-08 | (0, Inf) | 0.99 |
| Cisatracurium | 2.60 | (0.64, 10.65) | 0.18 | 0.19 | (0.002, 14.55) | 0.46 |
| Dexamethasone | 1.91 | (0.82, 4.88) | 0.13 | 8.35 | (0.42, 164.6) | 0.17 |
| Dexmedetomidine | 0.31 | (0.16, 0.63) | <0.01 | 0.17 | (0.03, 1.03) | 0.07 |
| Dobutamine | 0.19 | (0.06, 0.66) | <0.01 | 0.55 | (0.02, 17.36) | 0.73 |
| Dopamine | 0.28 | (0.12, 0.69) | <0.01 | 1.29 | (0.07, 24.57) | 0.87 |
| Ertapenem | 7.82 | (0.8, 76.27) | 0.07 | 1292.28 | (0.16, Inf) | 0.13 |
| Epinephrine | 0.46 | (0.22, 1.00) | 0.04 | 3.15 | (0.20, 48.55) | 0.41 |
| Hydromorphone | 0.62 | (0.34, 1.14) | 0.12 | 1.36 | (0.03, 53.87) | 0.87 |
| Lorazepam | 2.05 | (1.18, 3.61) | 0.01 | 2.67 | (0.45, 15.85) | 0.29 |
| Midazolam | 0.55 | (0.34, 0.91) | 0.01 | 0.49 | (0.08, 3.06) | 0.45 |
| Milrinone | 0.09 | (0.01, 0.68) | 0.01 | 2.56 | (0.02, 295.8) | 0.70 |
| Norepinephrine | 1.39 | (0.85, 2.28) | 0.18 | 2.83 | (0.42, 19.08) | 0.29 |
| Phenylephrine | 0.80 | (0.45, 1.45) | 0.46 | 2.42 | (0.53, 11.10) | 0.26 |
| Rocuronium | 0.50 | (0.27, 0.96) | 0.03 | 0.04 | (0.0004, 4.53) | 0.18 |
| Sodium Chloride | 0.60 | (0.34, 1.07) | 0.08 | 1.04 | (0.41, 2.63) | 0.93 |
| Vasopressin | 0.81 | (0.46, 1.46) | 0.49 | 1.19 | (0.22, 6.44) | 0.85 |
| Neuromuscular blockers | 0.59 | (0.33, 1.09) | 0.09 | 15.60 | (0.13, 1825) | 0.26 |
| Vasopressors | 1.18 | (0.73, 1.93) | 0.50 | 0.88 | (0.13, 6.20) | 0.90 |
| Steroids | 0.83 | (0.49, 1.43) | 0.50 | 0.46 | (0.007, 29.08) | 0.72 |
| Benzodiazepines | 0.90 | (0.55, 1.58) | 0.67 | 1.71 | (0.23, 12.60) | 0.60 |
| Inotropes | 0.41 | (0.21, 0.81) | <0.01 | 0.13 | (0.005, 3.45) | 0.23 |
| MRC-ICU at 24 hours of ICU admission | 0.98 | (0.96, 1.01) | 0.15 | 0.96 | (0.48, 1.92) | 0.91 |
| MRC-ICU at time of intubation | 0.97 | (0.96, 0.99) | <0.01 | 0.98 | (0.40, 2.43) | 0.97 |
| MRC-ICU at 24 hours after intubation | 1.00 | (0.99, 1.03) | 0.46 | 0.82 | (0.38, 1.78) | 0.62 |
| MRC-ICU at 24 hours after ICU admission (≥ 10) | 0.91 | (0.44, 1.88) | 0.79 | 2.36 | (0.47, 11.72) | 0.30 |
| MRC-ICU at time of intubation (≥10) | 0.97 | (0.54, 1.73) | 0.91 | 4.27 | (1.12, 16.33) | 0.04 |
| MRC-ICU at 24 hours after intubation (≥10) | 1.49 | (0.83, 2.7) | 0.18 | 1.21 | (0.29, 5.13) | 0.80 |
| **Flowsheet and laboratory values at time of intubation** | | | | | | |
| *Duration of support devices before intubation (HFNC, BiPAP, CPAP)* | | | | | | |
| 1 day | 0.47 | (0.22, 1.01) | 0.05 | - | - | - |
| 2 days | 2.1 | (0.12, 37.12) | 0.61 | 6.42 | (0.74, 55.59) | 0.11 |
| 3 days | 0.42 | (0.04, 4.09) | 0.45 | 12.19 | (0.75, 196.8) | 0.12 |
| ≥ 4 days | 3.5 | (0.69, 17.64) | 0.12 | 8.81 | (0.89, 86.86) | 0.09 |
| Elevated pulse n/N (%) | 1.3 | (0.57, 2.99) | 0.53 | 0.95 | (0.12, 7.45) | 0.96 |
| HCO_3_ < 20 mEq/L n/N (%) | 1.46 | (0.73, 2.96) | 0.28 | 0.71 | (0.21, 2.33) | 0.57 |
| PaCO_2_ | 0.94 | (0.91, 0.98) | <0.01 | 1.01 | (0.10, 10.22) | 0.99 |
| FiO_2_ | 1.02 | (1.01, 1.04) | <0.01 | 1.05 | (0.54, 2.07) | 0.88 |
| PaO_2_ / FiO_2_ | 1.00 | (0.99, 1.03) | 0.35 | 0.93 | (0.37, 2.30) | 0.88 |
| PEEP | 1.11 | (1.01, 1.23) | 0.03 | 0.74 | (0.37, 1.49) | 0.41 |
| Minute Ventilation | 0.98 | (0.9, 1.08) | 0.77 | 2.13 | (0.05, 97.07) | 0.71 |
| PaCO_2_ * Minute Ventilation | 0.99 | (1.00, 1.00) | 0.36 | 0.41 | (0.005, 36.41) | 0.71 |
| *Data are presented as n (%) or mean* ± *std unless otherwise stated.*  *SOFA: sequential organ failure assessment, APACHE II: Acute Physiology and Chronic Health Evaluation; ICU: intensive care unit; PEEP: positive end expiratory pressure* | | | | | | |

**Table E2.** Logistic regression for final model to predict prolonged mechanical ventilation

| **Variable** | **Odds Ratio** | **95% CI** | **p-value** |
| --- | --- | --- | --- |
| MRC-ICU at intubation | 0.75 | 0.55, 1.03 | 0.076 |
| Fluid overload before MV | 1.48 | 0.74, 3.40 | 0.351 |
| FiO_2_ | 1.14 | 0.85, 1.67 | 0.459 |
| PEEP | 1.19 | 0.79, 1.61 | 0.340 |
| PaO_2_ / FiO_2_ | 1.19 | 0.85, 1.81 | 0.320 |
| Pulse > 110 | 1.58 | 0.39, 2.24 | 0.310 |
| PaCO_2_ | 0.55 | 0.37, 0.83 | 0.005 |
| Minute Ventilation | 0.98 | 0.72, 1.33 | 0.888 |
| MRC-ICU: medication regimen complexity-intensive care unit; PEEP: positive end expiratory pressure; PaO_2_: partial pressure of oxygen; FiO_2_: fraction of inspired oxygen; PaCO_2_: partial pressure of carbon dioxide | | | |

**Figure E1.** The flowchart of model development.

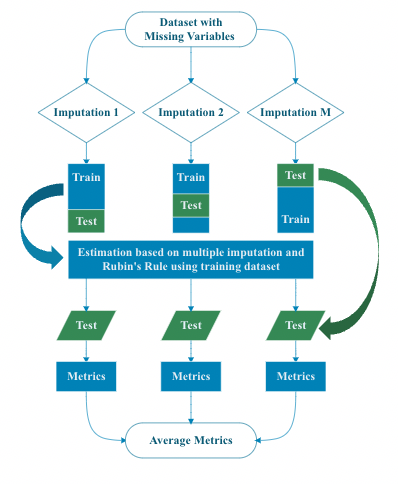

First, the dataset underwent 5 imputations to create 5 complete datasets. Each complete dataset was divided 85%/15% as training and testing sets. The estimation of training datasets was combined via Rubin’s Rule. Then, the model was applied to the test datasets and performance metrics were calculated. These were averaged and reported. Then, the dataset was imputed M times to result in M complete datasets. Each CC dataset was split into 5 folds with 1 fold saved testing, leaving the other 4 folds as training. Next, the imputed training datasets provided estimation with Rubin’s Rule as the normal estimation procedure to result in a combined estimation model. We then implemented prediction on each test dataset with this combined model. With the combined estimation results, predictions on each test dataset were completed and prediction metrics calculated. These results derived from M test datasets were averaged for the final evaluation. This process was adapted from R package ‘psfmi’ (https://missingdatasolutions.rbind.io/2021/02/mi-cross-validation/#methods, https://github.com/mwheymans/psfmi, accessed March 1, 2023).

**Figure E2.** Feature importance graph using Support Vector Machine

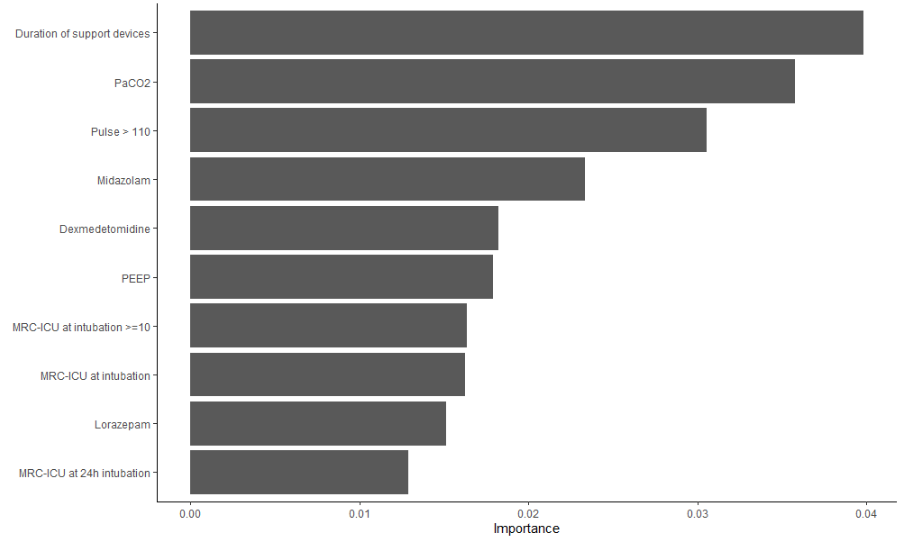

**Figure E3.** Feature importance graph using XGBoost

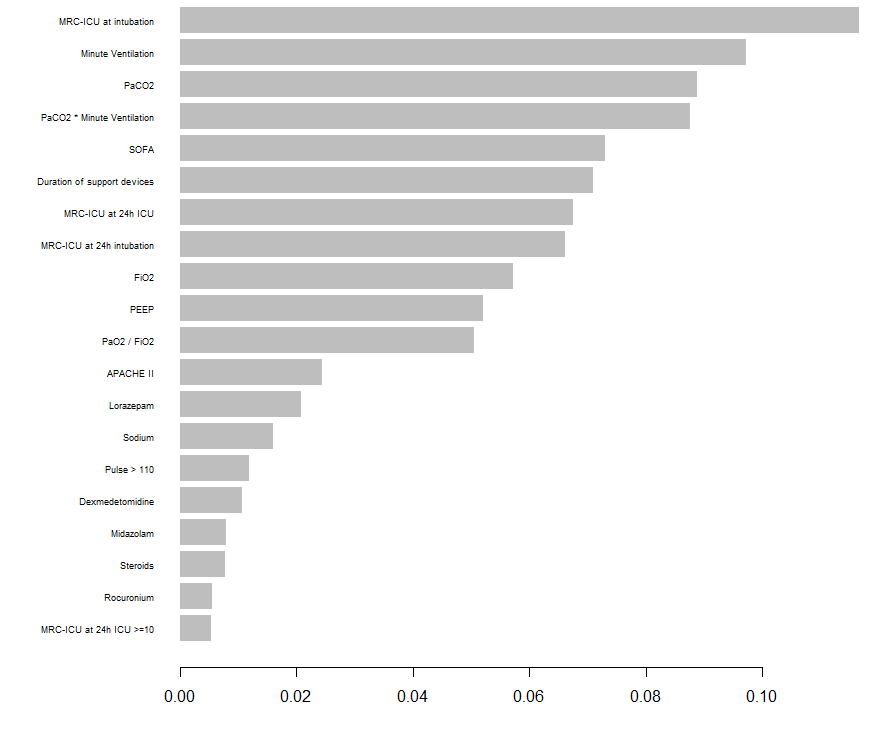
